# Intravenous Thrombolysis before Thrombectomy Improves Functional Outcome after Stroke Independent of Reperfusion Grade

**DOI:** 10.1101/2023.07.11.23292532

**Authors:** Annahita Sedghi, Daniel P. O. Kaiser, Ani Cuberi, Sonja Schreckenbauer, Claudia Wojciechowski, Ingeborg Friehs, Heinz Reichmann, Jessica Barlinn, Kristian Barlinn, Volker Puetz, Timo Siepmann

## Abstract

**Background:** We studied the effect of bridging intravenous thrombolysis (IVT) before thrombectomy for anterior circulation large vessel occlusion (acLVO) on functional outcome and scrutinized its dependence on grade of reperfusion and distal thrombus migration.

**Methods:** We included consecutive acLVO patients from our prospective registry of thrombectomy-eligible patients treated from 01/01/2017 to 01/01/2023 at a tertiary stroke center in Germany into a retrospective cohort study. We evaluated the effects of bridging IVT on functional outcome quantified via modified Rankin scale (mRS) at 90 days applying multivariable logistic and lasso regression including interaction terms with grade of reperfusion quantified via modified Thrombolysis in Cerebral Infarction (mTICI) scale and distal thrombus migration adjusted for demographic and cardiovascular risk profiles, clinical and imaging stroke characteristics, onset-to-recanalization time and distal thrombus migration. We performed sensitivity analysis using propensity score matching.

**Results:** In our study population of 1000 thrombectomy-eligible patients (513 females, median age 77 [67-84, interquartile range]) IVT emerged as predictor of favorable functional outcome (mRS 0-2) independent of mTICI (adjusted OR 0.49; 95% CI [0.32;0.75]; p=0.001). In those who underwent thrombectomy (n=812) the association of IVT and favorable functional outcome was reproduced (adjusted OR 0.49; 95% CI [0.31;0.74]; p=0.001) and was further confirmed on propensity score analysis where IVT led to a 0.35-point decrease in 90-day mRS score [ß=-0.35; 95CI% [-0.68;-0.01]; p=0.04]. The additive benefit of IVT remained independent of mTICI (ß=-1.79; 95% CI [-3.43;-0.15]; p=0.03) and distal thrombus migration (ß=-0.41; 95% CI [-0.69;-0.13]; p=0.004) on interaction analysis. Consequently, IVT showed an additive effect on functional outcome in the subpopulation of thrombectomy patients who achieved successful reperfusion (TICI≥2b; ß=-0.46; 95% CI [-0.74;-0.17]; p=0.002) and remained beneficial in those with unsuccessful reperfusion (TICI≤2a; ß=-0.47; 95% CI [-0.96;-0.01]; p=0.05).

**Conclusions:** In thrombectomy-eligible acLVO patients IVT improves functional outcome independent of grade of reperfusion and distal thrombus migration.

## Introduction

Intravenous thrombolysis (IVT) and thrombectomy improve clinical outcome of acute ischemic stroke (AIS) caused by cerebral anterior circulation large vessel occlusion (acLVO) in a highly time-dependent fashion with a rapid decline of efficacy with extending time from onset of symptoms.^1,2^ Several randomized controlled trials tested whether bridging IVT prior to thrombectomy has an additive beneficial effect on clinical outcome. These studies yielded conflicting results. While the European MR CLEAN NO IV trial and the Japanese SKIP trial failed to demonstrate non-inferiority of direct thrombectomy versus bridging IVT and thrombectomy, the Chinese studies DIRECT-MT and DEVT were able to show non-inferiority but were limited by wide margins.^3-6^ The latter observation was recently supported by a meta-analysis of clinical trials and observational studies that synthesized data from 36,123 patients and found slightly improved functional outcome and reperfusion rates in thrombectomy patients who also received bridging IVT.^7^ Moreover, the open-label, blinded-endpoint, randomized trials SWIFT-DIRECT and DIRECT-SAFE failed to show non-inferiority of omitting bridging IVT before thrombectomy and even found effect directions in favor of bridging IVT.^8,9^ Consequentially, the question whether bridging IVT adds value to thrombectomy beyond non-inferiority continues to be a topic of exploration. A recent retrospective cohort study in 746 acLVO patients who underwent thrombectomy but did not achieve successful reperfusion found improved functional outcome after 90 days possibly mediated by improved cerebral macrocirculation and microcirculation.^10^ Here, we aimed to assess if IVT in thrombectomy-eligible acLVO patients has a beneficial effect on functional outcome beyond an extent that can be explained by improvement of reperfusion as captured by modified Thrombolysis in Cerebral Infarction (mTICI) scale.

## Methods

### Study Design and Patients

We included patients from our prospective registry of consecutive potentially thrombectomy-eligible acLVO patients treated from 01/01/2017 to 01/01/2023 at the tertiary stroke center of University Hospital Carl Gustav Carus in Dresden, Germany into a retrospective cohort study. This study is reported in compliance with the Strengthening the Reporting of Observational Studies in Epidemiology (STROBE) statement.^11^ The STROBE checklist is shown in Supplemental Information S1. We included only adults who had acute ischemic stroke due to imaging-confirmed defined as occlusion of the intracranial segment of the internal carotid artery (ICA) or the M1 and/or M2 segment of the middle cerebral artery (MCA) with an established indication for thrombectomy. This encompassed those who have either undergone thrombectomy with or without prior IVT as well as those where thrombectomy was omitted after performance of IVT, e.g. because of early recanalization on repeated CT angiography or angiogram. We excluded patients with posterior circulation stroke as well as those with unknown onset of symptoms and unknown time of last seen well, unknown baseline National Institute of Health Stroke Scale (NIHSS) or missing modified Rankin Scale (mRS) score at 90 days. Details on study selection criteria and reasons for ommitance of thrombectomy are provided in the study flowchart (Figure 1).

**Figure 1:**
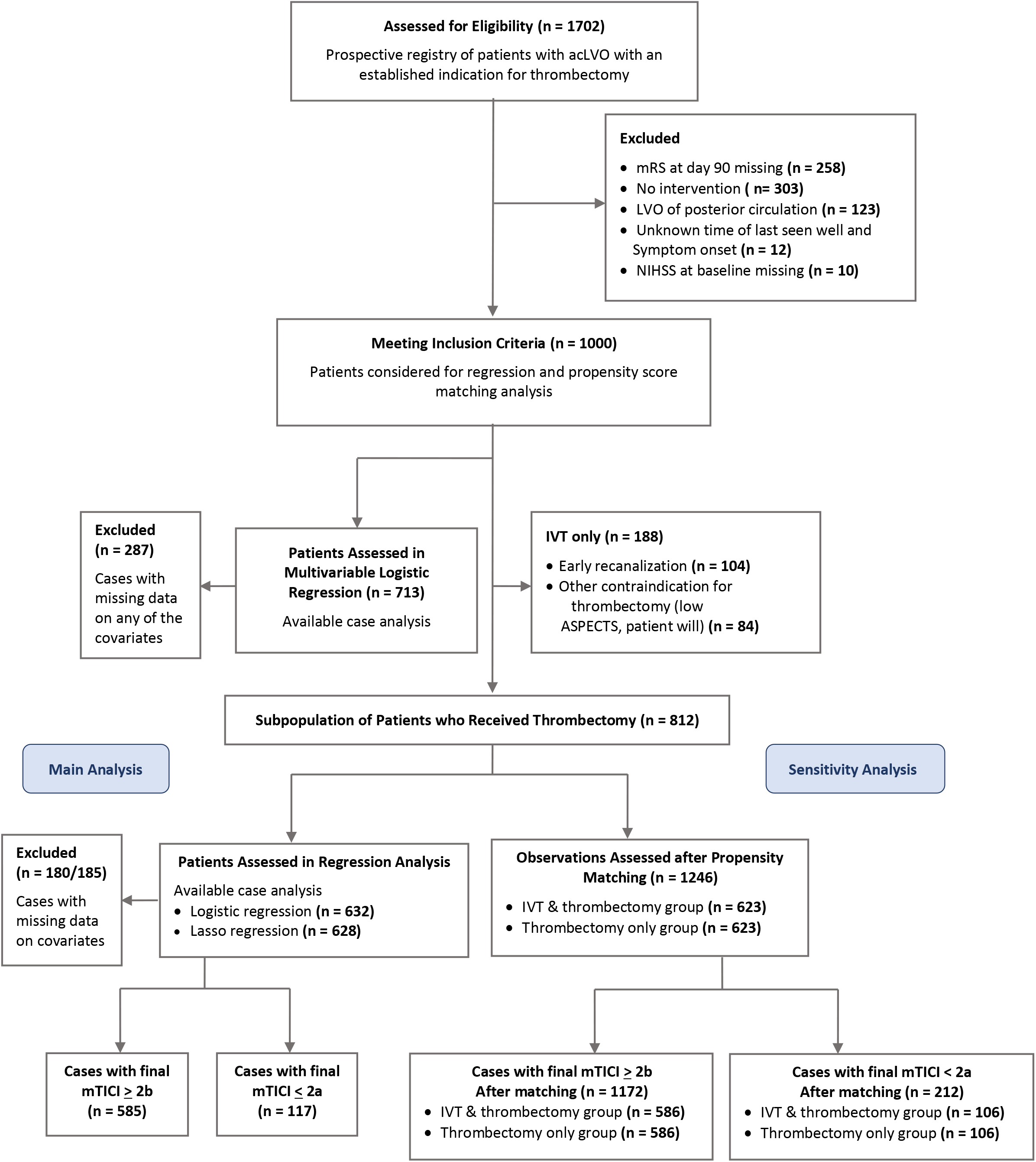
Study Flow Chart. Study flowchart illustrating the results of the screening and selecting patients for inclusion in the main analysis as well as the sensitivity and subgroup analyses of the study. acLVO, anterior circulation large vessel occlusion; mRS, modified Rankin scale; NIHSS, National Institutes of Health Stroke Scale; IVT, intravenous thrombolysis; ASPECTS, Alberta Stroke Program Early CT score; mTICI, modified treatment in cerebral infarction score.

### Clinical and Imaging Assessment

Our thrombectomy registry encompasses both mothership patients and drip-and-ship transfers from 13 community hospitals without a neurology department that are spokes of our telestroke network or from our eight partner hospitals who have neurological departments but no or limited thrombectomy capacity. Details of our regional stroke network have been published elsewhere.^12^ Our registry of thrombectomy-eligible patients comprises detailed data on demographic characteristics, premorbid condition, chronic comorbidity, cardiovascular risk profiles, medication, stroke etiology classified via Trial of Org 10172 in Acute Stroke Treatment (TOAST), neurological deficits rated via National Institute of Health Stroke Scale (NIHSS) by stroke physicians and functional outcome via modified Rankin Scale (mRS) scores at admission and discharge. Functional outcome was additionally obtained via a telephone interview 90 days after the day of intervention (IVT and/or thrombectomy) and favorable functional outcome was defined as a mRS of 0 to 2 at the time of this follow-up. Brain and vessel imaging findings included Alberta Stroke Program Early CT Score (ASPECTS) and mTICI, occlusion site and leptomeningeal collateral status. While mTICI was performed to quantify the grade of anterograde reperfusion of patent vasculature that supplies the target brain tissue after thrombus removal, the term recanalization is used henceforth to refer to the restoration of artery patency at the occlusion site.^13^ Treatment times were determined for onset-to-needle, onset-to-groin, onset-to-recanalization, needle-to-groin, needle-to-recanalization and groin-to-recanalization intervals.

Parameters of interest to our study that were not available in our registry were extracted via chart review by two independent investigators (AS, SS). A complete list of parameters and modes of their acquisition is provided in Table S1. We categorized vessel occlusion sites from proximal to distal into six groups as follows: (1.) tandem occlusion (extracranial ICA occlusion or high-grade stenosis preceding ipsilateral anterior circulation LVO); (2.) carotid-T occlusion (coexistence of distal intracranial ICA occlusion and ipsilateral proximal M1 and A1 occlusion); (3.) carotid-L occlusion (occlusion of distal intracranial ICA and proximal M1 segment); (4.) carotid-I occlusion (isolated intracranial ICA occlusion); (5.) isolated M1 occlusion; (6.) occlusion of M1/M2 junction or isolated M2 occlusion. We defined tandem occlusion as extracranial ICA occlusion or high-grade stenosis (≥ 70% North American Symptomatic Carotid Endarterectomy Trial stenosis, NASCET) preceding ipsilateral acLVO. Distal thrombus migration was defined as change from a proximal to distal category on angiogram compared to preceding CT angiography or on repeated CT angiography, e.g. following transfer from a drip-and-ship clinic to the mothership center. Distal thrombus migration beyond catheter accessibility resulting in omittance of thrombectomy or complete absence of vessel occlusion on repeated CT angiography or angiogram was considered early recanalization. In these patients, time of recanalization was defined as time of the first imaging (CT angiography or angiogram) that did not show a sustained occlusion within catheter reach. Thrombectomy with successful reperfusion was defined as a post-interventional mTICI score of 2b or higher. In cases without catheter angiography, e.g. because of early recanalization or insufficient core/penumbra mismatch on perfusion imaging at the mothership clinic following drip-and-ship transfer, mTICI scores were assessed using CT angiography post hoc by two experienced neuroradiologists (DK, AC) as previously described.^14^ Consensus was reached for ambiguous findings. Sedative regimen during thrombectomy was classified as general anesthesia or conscious sedation. Further details on the definitions of patient characteristics are shown in Supplemental Methods.

### Ethical Standard

Our study was approved by the local institutional review board (Ethikkommission an der TU Dresden, IRB reference number: EK 272072017). Written informed consent for participation was waived in accordance with the national legislation and the institutional requirements.

### Statistical Analysis

For analysis the study population was subdivided into three groups of patients receiving either IVT only, bridging IVT followed by thrombectomy or thrombectomy only. Independent continuous variables were checked for normality using descriptive and analytic (Shapiro-Wilk test) criteria. Between-group differences of demographic, clinical, imaging and procedural characteristics were assessed using Fisher’s exact test for binary data, Kruskal-Wallis test for categorical or non-normally distributed continuous data and one-way analysis of variance for normally distributed data where appropriate.

We performed multivariable regression to assess the association of IVT and its interaction with final mTICI score and distal thrombus migration with functional outcome at 90 days in the entire study population. Covariates adjusted for where chosen by clinical reasoning and comprised age, premorbid dependency, chronic disease possibly impairing functional independence, malignancy, arterial hypertension, HbA1c (%), low-density lipoprotein (LDL, mg/dl), smoking, stroke etiology, NIHSS at baseline, Alberta Stroke Program Early CT score (ASPECTS), vessel site, tandem occlusion, carotid T occlusion, mTICI score, emergency carotid stenting, thrombectomy, onset-to-recanalization time, and stroke etiology as defined by TOAST category. Further definitions of covariates are detailed in Supplemental Methods. Multivariable regression was repeated in the subgroup of patients who have undergone thrombectomy with and without preceding IVT. Residuals were tested for normality. Multicollinearity was assessed by calculating the variable inflation factor (VIF) for all covariates in the regression model. A VIF value of 1 indicates no multicollinearity, whereas VIF values above 5 indicate relevant multicollinearity. Where multicollinearity impaired interpretability of the regression model, double selection lasso linear regression for inference using cross-validation and controlling for all covariates included in the original regression model was utilized to obtain reliable results. Interaction terms were included in regression models to assess independency of effects of IVT, distal thrombus migration and final mTICI score on functional outcome after 90 days.

We conducted a sensitivity analysis using propensity score matching to test the robustness of the results on the average effect of IVT on 90-day functional outcome in the subpopulation of patients that underwent thrombectomy while additionally accounting for the non-randomized study design. Propensity scores were estimated by multivariable logistic regression incorporating the same covariates as used in the main model with the addition of sedative regimen applied during thrombectomy. The maximum allowed difference in propensity scores for matching (caliper value) was targeted to be ≤0.2. Standardized differences and variance ratios were calculated to assess balance of covariates between the two groups of patients receiving either bridging IVT followed by thrombectomy or thrombectomy alone. We aimed for a standardized difference of 0±0.1 and a variance ratio of 1±0.25. The aforementioned analyses were repeated in subgroups of patients who underwent thrombectomy with and without successful reperfusion. Significance level was set at α = 0.05. Available case analysis was performed. The number of missing registry data is reported in Table S2 and was low. All analyses were performed using the statistical software package Stata® (StataCorp. 2021. Stata Statistical Software: Release 17. College Station, TX: StataCorp LLC).

## Results

### Study Population

We included 1000 patients with AIS due to acLVO (513 females; median age 77 years [interquartile range, IQR 67-84 years]; baseline NIHSS 15 [IQR, 11-19]; baseline mRS 5 [4-5]; median ASPECTS 7 [IQR, 6-9]; median onset-to-needle time 106 minutes [IQR, 81-135 minutes]; median onset-to-recanalization time 287 minutes [IQR, 219-358 minutes]), of whom 402 (40.2%) received bridging IVT before thrombectomy, 188 (18.8%) received only IVT and 410 (41.0%) underwent thrombectomy alone. Ninety-day mRS was 4 [IQR, 1;6] in patients who received only IVT, 3 [IQR, 1;5] in those who received bridging IVT and subsequent thrombectomy and 4 [IQR, 2;6] those who underwent only thrombectomy. Out of all patients who received IVT (n=590), 112 (18.9%) patients showed early recanalization. Distal thrombus migration was observed in 40 (6.7%) patients. Early recanalization occurred in 20 (12.6%) of patients who received IVT following direct transportation to the mothership hospital of our telestroke network (n=159) versus 92 (21.4%) of those who received IVT at a drip-and-ship hospital (n=431). Distal thrombus migration occurred in 7 (4.4 %) of patients who received IVT following direct transportation to the mothership hospital of our telestroke network (n=159) versus 33 (7.6%) of those who received IVT at a drip-and-ship hospital (n=431). Demographic features, vascular risk profiles as well as clinical and imaging characteristics are detailed in Table 1.

**Table 1:** Demographic and Baseline Characteristics.

|  | IVT (n= 188) | IVT+MT (n=402) | MT (n=410) | p-value |
| --- | --- | --- | --- | --- |
| **Demographic characteristics** |  |  |  |  |
| Age, years (median [IQR]) | 77 [68;84] | 76 [64;83] | 79 [69;84] | 0.013 |
| Sex, female (n, %) | 94, 49.5 | 211, 52.8 | 208, 50.7 | 0.628 |
| Premorbid condition |  |  |  | 0.008 |
| Independent (n, %) | 135, 74.6 | 310, 77.1 | 277, 67.6 |  |
| Needs assistance (n, %) | 46, 25.1 | 92, 22.9 | 133, 32.4 |  |
| Chronic disease (n, %) |  |  |  | 0.076 |
| No organ system | 107, 58.8 | 262, 65.3 | 239, 58.3 |  |
| 1 Organ system | 52, 28.6 | 101, 25.2 | 120, 29.3 |  |
| 2 Organ systems | 20, 11.0 | 35, 8.7 | 45, 11.0 |  |
| 3 Organ systems | 3, 1.7 | 3, 0.8 | 6, 1.5 |  |
| 4 Organ systems | - | - | - |  |
| Malignancy (n, %) |  |  |  | 0.0003 |
| None | 165, 87.7 | 361, 89.8 | 328, 80.0 |  |
| 1 organ system | 20, 10.6 | 39, 9.7 | 79, 19.3 |  |
| 2 organ systems | 3, 1.6 | 2, 0.5 | 2, 0.5 |  |
| 3 organ systems | - | - | 1, 0.2 |  |
| **Cardiovascular risk factors** |  |  |  |  |
| Arterial hypertension (n, %) | 159, 88.3 | 352, 88.0 | 369, 90.0 | 0.634 |
| Diabetes mellitus (n, %) | 54, 30.3 | 112, 27.9 | 120, 29.3 | 0.810 |
| HbA1c, % (median [IQR]) | 5.7 [5.4;6.2] | 5.8 [5.5;6.2] | 5.8 [5.4;6.3] | 0.910 |
| LDL, mg/dl (mean, SD) | 2.7 ± 1.0 | 2.5 ± 1.0 | 2.3 ± 0.9 | 0.244 |
| **Preventive pharmacological treatment** |  |  |  |  |
| Statin (n, %) | 56, 31.6 | 123, 30.8 | 141, 34.4 | 0.530 |
| Antiplatelet therapy (n, %) | 62, 35.6 | 124, 31.4 | 82, 20.1 | 0.000 |
| **Stroke characteristics** |  |  |  |  |
| Wake-up | 13, 7.0 | 33, 8.2 | 130, 32.9 | 0.000 |
| NIHSS at baseline (median, [IQR]) | 16 [12;20] | 16 [11;19] | 15 [10;19] | 0.026 |
| NIHSS at discharge (median, [IQR]) | 9 [2;42] | 5 [1;15] | 10 [3;19] | 0.0001 |
| mRS at baseline (median [IQR]) | 5 [4;5] | 5 [4;5] | 5 [4;5] | 0.492 |
| mRS at discharge (median [IQR]) | 4 [2;6] | 3 [2;5] | 4 [3;5] | 0.0001 |
| mRS at 90 days (median [IQR]) | 4 [1;6] | 3 [1;5] | 4 [2;6] | 0.0001 |
| mRS (0-2) at 90 days (n, %) | 63, 33.5 | 184, 45.8 | 304, 74.2 | 0.000 |
| ASPECTS (median [IQR]) | 7 [4;9] | 8 [6;9] | 7 [6;9] | 0.003 |
| Occlusion side, left (n, %) | 93, 49.5 | 212, 52.8 | 198, 48.3 | 0.434 |
| Occlusion site (n, %) |  |  |  | 0.147 |
| ICA intracranial | 7, 3.7 | 7, 1.7 | 2, 0.5 |  |
| L, M1 prox.-distal | 142, 75.5 | 335, 83.3 | 329, 80.2 |  |
| M1/2-transition, M2 prox.- distal | 39, 20.7 | 60, 14.9 | 79, 19.3 |  |
| Carotid T occlusion (n, %) | 24, 12.8 | 36, 8.9 | 44, 10.8 | 0.352 |
| Tandem occlusion (n, %) | 32, 17.2 | 69, 17.2 | 36, 8.8 | 0.001 |
| Leptomeningeal collaterals on DSA (n, %) | - | 320, 81.0 | 354, 86.6 | 0.025 |
| **TOAST classification (n, %)** |  |  |  | 0.299 |
| Large artery atherosclerosis | 29, 16.1 | 84, 20.9 | 66, 16.2 |  |
| Cardioembolism | 104, 57.8 | 229, 57.0 | 247, 60.7 |  |
| Small vessel occlusion | 1, 0.6 | - | - |  |
| Stroke of other determined etiology | 3, 1.7 | 13, 3.2 | 10, 2.5 |  |
| Stroke of undetermined etiology | 43, 23.9 | 76, 18.9 | 84, 20.6 |  |
| **Interventions (n, %)** |  |  |  |  |
| Drip and ship | 157, 83.5 | 274, 68.2 | 255, 62.2 | 0.000 |
| Carotid stent (emergency) | 2, 1.1 | 49, 12.2 | 38, 9.3 | 0.000 |
| **Sedative regimen (n, %)** |  |  |  | 0.430 |
| Conscious sedation | - | 115, 29.6 | 117, 28.8 |  |
| General anesthesia | - | 273, 70.4 | 289, 71.2 |  |
| **Procedural times, min (median [IQR])** |  |  |  |  |
| Onset unclear | 5, 2.7 | 11, 2.7 | 112, 27.3 | 0.000 |
| Onset-to-needle | 108 [82;135] | 105 [80;135] | - | 0.560 |
| Onset-to-groin | 165 [130;210] | 240 [165;285] | 240 [175;313] | 0.021 |
| Onset-to-recanalization | 210 [171;250] | 302 [231.5;354.5] | 296 [237;385] | 0.0001 |
| Groin-to-recanalization | - | 52 [33;83] | 53 [33;88] | - |
| Needle-to-groin | 60 [42;129] | 128 [60;160] | - | 0.020 |
| **Procedural outcomes (n, %)** |  |  |  |  |
| Early recanalization | 104, 55.3 | 8, 2.0 | - | - |
| Distal thrombus migration | 4, 2.1 | 36, 9.0 | 3, 0.7 | 0.000 |
| Thrombectomy frustrate | - | 37, 9.4 | 53, 13.0 | 0.232 |
| mTICI (n, %) |  |  |  | 0.0001 |
| 0 | 69, 37.5 | 30, 7.5 | 42, 10.4 |  |
| 1 | 5, 2.7 | 2, 0.5 | 6, 1.5 |  |
| 2a | 7, 3.8 | 17, 4.2 | 23, 5.6 |  |
| 2b | 42, 22.8 | 137, 34.1 | 139, 33.9 |  |
| 2c, 3 | 61, 33.2 | 216, 53.7 | 200, 48.8 |  |
| Successful reperfusion (mTICI ≥ 2b) | 107, 56.9 | 353, 87.8 | 339, 82.7 | 0.000 |

IVT, intravenous thrombolysis; MT, mechanical thrombectomy; IQR, interquartile range; SD standard deviation; LDL, low density lipoprotein; HbA1C, Haemoglobin A1c; NIHSS, National Institutes of Health Stroke Scale; mRS, modified Rankin scale; ASPECTS, Alberta Stroke Program Early CT score; ICA, internal carotid artery; DSA, digital subtraction angiography; mTICI, modified treatment in cerebral infarction score

### Association of IVT and Favorable Functional Outcome in Thrombectomy-Eligible Patients

In the entire study population multivariable linear regression substantiated a positive predictive association between performance of IVT and favorable functional outcome independent of grade of reperfusion quantified via mTICI and onset-to-recanalization time with additional adjustment for all predefined clinically relevant covariates (OR 0.49; 95% CI [0.32;0.75]; p=0.001). This positive association remained significant when we included only patients who received bridging IVT with subsequent thrombectomy (OR 0.49; 95% CI [0.32;0.74]; p=0.001). Besides, ASEPCTS was positively associated with favorable functional outcome, whereas negative predictive associations with favorable functional outcome were noted for premorbid dependency, presence of tandem occlusion, history of malignancy, higher baseline NIHSS score, age, higher HbA1c and longer onset-to-recanalization time as detailed in Table S3.

We were able to confirm a positive independent association on sensitivity analysis using propensity score matching. Here, IVT was associated with an average 0.35-point decrease in the mRS score at day 90 in patients who received bridging IVT compared to patients who received thrombectomy alone (ß=-0.35; 95% CI [-0.68;-0.01]; p=0.04). The standardized differences and variance ratios are displayed in Table S4 and indicate a good match.

### Modulation of the Effect of IVT on Functional Outcome by Grade of Reperfusion and Distal Thrombus Migration

We went on to assess whether the observed beneficial main effect of bridging IVT on functional outcome might be modulated by the interaction with grade of reperfusion. In the post estimation analysis, we observed multicollinearity for the effects of bridging IVT, grade of reperfusion and their interaction possibly undermining a significant contribution of bridging IVT and mTICI score to 90-day functional outcome (mean VIF 19.8). On lasso regression for inference the beneficial main effect of bridging IVT on 90-day functional outcome persisted after accounting for its interaction with grade of reperfusion (ß=-1.79; 95% CI [-3.43;-0.15]; p=0.03). The interaction effect was non-significant (p>0.05) as detailed in Table S5. Higher mTICI scores were incrementally associated with improvement of 90-day functional outcome (mTICI2a: ß=-0.85; 95% CI [-1.63; -0.07]; p=0.03. mTICI 2b: ß=-1.57; 95% CI [-2.19; -0.96]; p=<0.001. mTICI 2c/3: ß=-1.79; 95% CI [-2.42; -1.16]; p=<0.001) without compromising the independent beneficial effect of IVT on 90-day functional outcome. Bridging IVT was also associated with an improved 90-day functional outcome when accounting for its interaction with distal thrombus migration (ß=-0.41; 95% CI [-0.69; -0.12]; p=0.004). The interaction of the effect of IVT on 90-day functional outcome with distal thrombus migration was negative (ß=0.34; 95% CI [-1.7; 0.49] p=0.42).

### Effects of Bridging IVT on Functional Outcome Following Thrombectomy with Successful Reperfusion

In due consideration of the aforementioned absence of an interaction between grade of reperfusion and effect of bridging IVT on functional outcome, we performed complementary subgroup analyses in patients who underwent thrombectomy with and without successful reperfusion defined as a final mTICI score of ≥ 2b and ≤ 2a, respectively. In patients with successful reperfusion bridging IVT remained a positive predictor for improved 90-day functional outcome on multivariable linear regression adjusted for age, sex, premorbid condition, baseline NIHSS, ASPECTS, vessel occlusion site, thrombus migration, presence of tandem occlusion or carotid T occlusion, onset-to-recanalization time, carotid stenting, chronic disease, malignancy, arterial hypertension, HbA1c, LDL, TOAST category, presence of leptomeningeal collaterals and sedative regimen (ß=-0.45; 95% CI [-0.74;-0.17]; p=0.002). A list of covariates associated with an improved 90-day functional outcome is detailed in Table S6. We were able to reproduce the beneficial effect of bridging IVT on 90-day functional outcome in thrombectomy with successful reperfusion on propensity score matching analysis where bridging IVT was associated with an 0.50-point decrease in final mRS score (ß=-0.50; 95% CI [- 0.84;-0.16]; p=0.004). The standardized differences and variance ratios are displayed in Table S6 and overall indicate a good match.

### Effects of Bridging IVT on Functional Outcome in Thrombectomy with Unsuccessful Reperfusion

In the subgroup of thrombectomy patients who did not show successful reperfusion (final mTICI ≤ 2a) bridging IVT was still associated with improved 90-day functional outcome on multivariable regression (ß=-0.47; 95% CI [-096;0.009]; p=0.05). A reduced number of clinically reasonable covariates was included in the multivariable linear regression model to avoid overfitting due to the lower sample size of this subgroup. The covariates were age, premorbid condition, baseline NIHSS, ASPECTS, vessel occlusion site, presence of tandem occlusion and TOAST category. Covariates significantly modifying functional outcome are detailed in Table S8.

Standardized differences and variance ratios of the sensitivity analysis using propensity matching are displayed in Table S9 and overall indicate a good match. Sensitivity analysis confirmed a beneficial association of bridging IVT prior to thrombectomy with 90-day functional outcome yielding a 0.37-point increase in 90-day mRS (ß=-0.37; 95% CI [-0.74;-0.01]; p=0.05) when compared with thrombectomy alone.

## Discussion

The main finding of this study is that bridging IVT for anterior circulation stroke due to acLVO compared to thrombectomy alone improves functional outcome to an extent that cannot be explained solely by facilitated reperfusion as quantified by mTICI score.

In the light of inconclusive observations on the effect of bridging IVT for acLVO on functional outcome from several RCTs and meta-analyses^3-9,15^, recent research focused on deeper exploration of patient and treatment-related characteristics that might modulate this effect. A retrospective analysis of the International Stroke Perfusion Imaging Registry (n=323) suggested that bridging IVT (n=241) is beneficial in patients with fast growing infarct core due to more rapid completion of thrombectomy resulting in a reduced final size of the ischemic lesion.^16^ In a pre-specified secondary analysis of the DIRECT MT trial (n=640) functional outcome was overall worse in more proximal compared to more distal occlusions but the effect of bridging IVT (n=325) on functional outcome was not modulated by occlusion site when differentiating ICA, M1 and M2 occlusions.^17^ In posthoc analyses of the prospective observational studies INTERRSeCT and MR CLEAN registry distal thrombus migration on repeated CTA or angiogram was associated with better functional outcome acLVO patients.^18,19^ In this regard another posthoc analysis of DIRECT MT found that distal thrombus migration resulting in an eTICI Score ≥ 2a prior to thrombectomy, correlated with an improved functional outcome regardless of the final eTICI score, leading the authors to conclude that the use of bridging IVT, which promotes early reperfusion, reduces ischemia time in the reperfused tissue and allows for blood flow through already recanalized collateral vessels.^20^ In our study, the observed net effect of IVT on functional outcome was adjusted for onset-to-recanalization time to account for cases of early recanalization following IVT with subsequent ommitance of thrombectomy in the overall population of acLVO patients as well as for procedure times in the subpopulation of patients who received thrombectomy. Furthermore, in our study distal thrombus migration and early recanalization could be detected in 152 (25.8 %) of the follow-up CT angiographies or angiorams in patients, who received IVT (n=590). The fraction was higher in patients who received IVT at the drip-and-ship hospital compared with the mothership clinic (n=92+33=125 von 431, 29% vs. n=20+7=27 von 159, 17.0%) likely due to the dilutive effects of a longer exposure times to IVT as previously suggested.^17-20^ Nevertheless, the positive net effect of bridging IVT on functional outcome was modulated neither by distal thrombus migration nor by the final mTICI score. Since these observations suggest that the success of bridging IVT does not depend solely on the success of subsequent thrombectomy, we repeated analysis in subgroups of patients who underwent thrombectomy with and without successful reperfusion. In both subgroups we were able to confirm a positive net effect of bridging IVT on improved functional outcome on repeated primary analysis as well as sensitivity analysis. Our observation in the subgroup of thrombectomy patients not achieving successful reperfusion interventionally is consistent with a recent cohort study (n=756) showing improved functional outcome in acLVO patients with unsuccessful reperfusion following thrombectomy.^10^ A continued beneficial effect of IVT beyond the physiologically momentous event of recanalization during thrombectomy might be explained by a sustained pharmacologic effect of recombinant tissue-type plasminogen activator (rtPA) on the cerebral microcirculation. Congruently, in a transient middle cerebral artery occlusion rat model, rtPA improved microvascular perfusion by reducing platelet aggregation in a fibrinogen-dependent fashion with consequential reduction of downstream microvascular thrombosis.^22^ Translating this observation into human stroke survivors, a randomized placebo-controlled clinical trial in 121 acLVO patients found that intra-arterial application of rtPA in patients after thrombectomy with successful reperfusion increased the likelihood of achieving an excellent functional outcome at 90 days defined as an mRS score of 0-1.^23^

### Strengths and Limitations

Our observation of improved functional outcome following bridging IVT derived from a retrospective analysis of a prospective registry of thrombectomy-eligible patients with partially imbalanced groups but showed high reproducibility on sensitivity analysis using propensity score matching and is independent of the grade of reperfusion. While the direction and significance of the assertions in our analyses are meaningful and unambiguous, the exact values of the continuous coefficients may be of limited absolute interpretability due to varying ranges of the different parameters by nature. Despite the goodness of fit of our regression models that is considered decent, still some amount of data remains unexplained by multivariable regression models. Real-world big data analysis could help identify new parameters that modulate the effect of bridging IVT while avoiding overfitting. A major reason why the question whether bridging IVT before thrombectomy is beneficial has not yet been answered indubitably by observational or interventional research, including our data, might be that regression analysis with multiple covariates is utilized as tacit predictive model for which studies have not been powered for sufficiently. While our registry of acLVO patients requiring thrombectomy is of multicentric nature, encompassing a large telestroke network, thrombectomy was solely performed at the mothership clinic. However, highly standardized acute and post-interventional stroke care as well as reproducibility of observations on propensity score-based sensitivity analyses and subgroup analyses support the internal validity and generalizability of our findings.

### Conclusion

Bridging IVT is beneficial with respect to functional outcome in patients with acLVO stroke who have an established indication for thrombectomy regardless of the achieved grade of reperfusion as quantified by the mTICI score and occurrence of distal thrombus migration.

## Data Availability

The data that support the findings of this study are available from the corresponding author, TS, upon reasonable request.

## Non-Standard Abbreviations and Acronyms

IVT: Intravenous thrombolysis
AIS: acute ischemic stroke
acLVO: anterior circulation large vessel occlusion
mTICI: modified Thrombolysis in Cerebral Infarction
STROBE: Strengthening the Reporting of Observational Studies in Epidemiology
MCA: middle cerebral artery
NIHSS: National Institute of Health Stroke Scale
mRS: modified Rankin Scale
TOAST: Trial of Org 10172 in Acute Stroke Treatment
ASPECTS: Alberta Stroke Program Early CT Score
VIF: variable inflation factor
rtPA: recombinant tissue-type plasminogen activator

## Acknowledgments

Daniel P. O. Kaiser is supported by the Joachim Herz Foundation. Timo Siepmann received grants from the German Federal Ministry of Health and Kurt Goldstein Institut, royalties from Astrazeneca for consulting and from Dresden International University for serving as a program director and a lecturer of the Master’s Program in Clinical Research. None of these activities were related to the current study.

## Supplemental Material

Supplemental Methods

Tables S1-S9

STROBE checklist

Reference S1

